# Nicotine and tobacco product use among US middle and high school students, 2024-25

**DOI:** 10.64898/2026.03.20.26348931

**Authors:** Haoxiong Sun, Yu Jiang, Harry Tattan-Birch, Siyuan Fan, Sharon Cox, Sarah E. Jackson

## Abstract

**Importance:** The overall prevalence of youth nicotine and tobacco product use has declined over recent years, but the product landscape continues to evolve rapidly, particularly with new disposable e-cigarettes and oral nicotine pouches.

**Objective:** To examine changes between 2024 and 2025 in the prevalence of nicotine and tobacco product use among US middle and high school students and describe shifts in product characteristics among current e-cigarette and nicotine pouch users.

**Design, Setting, and Participants:** Repeated cross-sectional study using nationally representative data from the 2024 and 2025 National Youth Tobacco Survey (NYTS), a school-based survey of US students in grades 6–12 (approximately ages 11-18). The analytic sample included 29,678 students in 2024 and 23,557 students in 2025.

**Exposures:** Survey year (2025 vs 2024).

**Main Outcomes and Measures:** Past 30-day use of nicotine/tobacco products, including e-cigarettes, nicotine pouches, cigarettes, and other combustible and non-combustible products. Among current e-cigarette and nicotine pouch users, frequency of use, device type, brands, and flavors were assessed.

**Results:** In 2025, 7.2% (95% CI, 6.4-8.2%) of US middle and high school students reported past 30-day use of any nicotine/tobacco product, compared with 8.1% (7.4-8.9%) in 2024. E-cigarettes remained the most commonly used product (5.2%, 4.5-5.9%); 1.7% (1.4-2.1%) used nicotine pouches, 1.7% (1.4-1.9%) smoked cigarettes, and 2.7% (2.4-3.1%) smoked any combustible tobacco product. Among current e-cigarette users, 40.7% (36.7-44.9%) reported frequent use and 27.0% (24.0-30.2%) reported daily use in 2025. Disposable e-cigarette use increased from 55.8% (52.6-59.0%) in 2024 to 66.7% (62.5-70.7%) in 2025, while pod/cartridge device use declined. Flavored product use was reported by 90.0% of e-cigarette users and 88.0% of nicotine pouch users. The most commonly reported brands were Geek Bar among e-cigarette users (61.1%) and ZYN among nicotine pouch users (69.4%).

**Conclusions and Relevance:** Overall youth nicotine and tobacco use remains relatively low, but the product landscape is evolving rapidly, with increasing disposable device use and shifting brand preferences. These findings highlight the importance of ongoing, product-specific surveillance to inform public health strategies and regulatory policies.

## Introduction

Although cigarette smoking has decreased steadily among US youth since the mid-1990s (1), adolescent use of nicotine and tobacco products remains an important focus for public health surveillance. These products include cigarettes and other combustible tobacco products, heated tobacco products, and non-combustible products such as e-cigarettes, smokeless tobacco and, more recently, nicotine pouches. The marketplace is evolving rapidly, with changes in product availability, device types, brand popularity, and flavors. Alongside ongoing regulatory actions targeting flavored products and premarket authorization requirements, these shifts underscore the need for continued monitoring to better understand youth nicotine use patterns and inform public health and regulatory responses.

The National Youth Tobacco Survey (NYTS) provides nationally representative data used by the Centers for Disease Control and Prevention (CDC) and the US Food and Drug Administration (FDA) to monitor youth nicotine and tobacco product use in the US (2–5). These data enable timely assessment of changes in product prevalence as well as characteristics such as frequency of use, device type, brands, and flavors among youth users. The primary aim of this study was to examine changes between 2024 and 2025 in the prevalence of nicotine and tobacco product use among US middle and high school students. A secondary aim was to describe changes in frequency of use, brand, device type, and flavor preferences among current e-cigarette and nicotine pouch users.

## Methods

### Study Design and Participants

This repeated cross-sectional study used publicly available, de-identified data from the 2024 and 2025 National Youth Tobacco Survey (NYTS) and therefore did not require institutional review board approval. The study followed the Strengthening the Reporting of Observational Studies in Epidemiology (STROBE) reporting guideline for cross-sectional studies.

The NYTS is a school-based, self-administered, web-based survey of U.S. middle school (grades 6-8) and high school (grades 9-12) students (approximately ages 11-18) conducted by the Centers for Disease Control and Prevention and the U.S. Food and Drug Administration (6). The survey uses a stratified, three-stage cluster sampling design to generate nationally representative estimates for students attending public and private schools in the 50 states and the District of Columbia (6).

Data were collected during January through May in both survey years. In 2024, 29,861 students from 283 schools participated, with an overall response rate of 33.4% (5). In 2025, 23,630 students participated, with a response rate of 29.7% (6). After excluding respondents with missing information on their school level, the analytic sample comprised 29,678 students in 2024 and 23,557 students in 2025.

### Measures

Full details of survey measures are provided in the NYTS methodology report (6, 7). Key measures are summarized below.

#### Nicotine/tobacco product use

Past 30-day (current) use was defined as use on 1 or more days during the 30 days preceding the survey. Current use was assessed for 12 nicotine/tobacco products: e-cigarettes, nicotine pouches, manufactured cigarettes, roll-your-own cigarettes, cigars (including cigarillos and little cigars), smokeless tobacco (including chewing tobacco, snuff, and dip), snus, hookahs, heated tobacco products, oral nicotine products (including lozenges, discs, tablets, gums, and dissolvable tobacco products), pipe tobacco, and bidis.

Three composite measures were constructed: (1) any nicotine/tobacco product, defined as current use of at least 1 of the 12 products; (2) any combustible tobacco product, defined as current use of manufactured cigarettes, roll-your-own cigarettes, cigars, hookahs, pipe tobacco, or bidis; and (3) cigarettes, defined as current use of manufactured or roll-your-own cigarettes.

#### E-cigarette and nicotine pouch use characteristics

Students who currently use e-cigarettes or nicotine pouches reported their frequency of use (number of days used in the past 30 days), brands used, and flavors used. Four categories were created to characterize frequency of use: occasional (1–5 days), moderate (6–19 days), frequent (≥20 days), and daily (all 30 days) (8). Current e-cigarette users also reported the device type they used most often (disposable, pod or cartridge-based, or tank or mod system).

For check-all-that-apply brand and flavor items, each response option was coded as a separate binary indicator. Two composite flavor variables were created for both e-cigarettes and nicotine pouches: mint/menthol (any reported use of mint or menthol) and any flavored (use of at least 1 flavor other than tobacco or unflavored). Because some brand response options differed between survey years, cross-year brand comparisons were interpreted cautiously.

#### Demographic characteristics

Demographic characteristics included sex (female, male), school level (middle school, high school), and race and ethnicity. Race/ethnicity were self-reported and categorized as Hispanic, non-Hispanic American Indian or Alaska Native, non-Hispanic Asian, non-Hispanic Black, non-Hispanic White, and non-Hispanic multiracial or other.

### Statistical Analysis

All analyses accounted for the NYTS complex survey design by incorporating primary sampling units, strata, and final analytic weights to produce nationally representative estimates and appropriate variance estimates.

For the 2024 and 2025 middle school subgroup, two variance estimation strata contained singleton or near-singleton primary sampling units, producing non-estimable confidence intervals (CIs). To address this, adjacent strata were collapsed before variance estimation, and sensitivity analyses using alternative variance estimation methods confirmed the robustness of these estimates **in Supplementary Table 1**(9, 10).

Weighted prevalence estimates and 95% CIs were calculated for past-30-day (current) use of each product by year, overall and stratified by school level and sex (sex-stratified results are provided in **Supplementary Table 2**). CIs were estimated using the logit transformation method. National estimated numbers of users were calculated from survey-weighted population totals and rounded to the nearest 10,000.

Among current e-cigarette users and nicotine pouch users, weighted proportions and 95% confidence intervals were estimated for frequency of use, device type, brands, and flavors. Year-to-year differences in weighted proportions were assessed using design-based Wald tests. For comparisons of use characteristics among current users, unadjusted differences in weighted proportions and corresponding Wald test P values are reported, as these analyses describe within-user characteristics rather than population-level prevalence.

Changes in prevalence between 2024 and 2025 were assessed using Wald tests based on the pooled standard error of the difference in weighted proportions. Adjusted prevalence ratios (aPRs) and 95% confidence intervals were estimated using survey-weighted modified Poisson regression with a log link and design-based variance estimation. In overall models, the main exposure was survey year (2025 vs 2024), and models adjusted for school level, sex, and race/ethnicity. School-level subgroup models adjusted for sex and race/ethnicity.

All statistical tests were 2-sided. For each analysis, respondents with missing data on the outcome or model covariates were excluded from that specific analysis (complete-case analysis). Analyses were conducted in R, version 4.5.2 (R Foundation for Statistical Computing).

## Results

### Past 30-Day Use of Any Nicotine/Tobacco Product

In 2025, 7.2% of U.S. middle and high school students reported past-30-day use of any nicotine/tobacco product, corresponding to approximately 2.0 million students, compared with 8.1% in 2024 (∼2.23 million) (**Table 1**). Adjusted analyses indicated lower prevalence in 2025 than in 2024, although the confidence interval for the change included the null value (aPR 0.89; 95% CI 0.77–1.02).

**Table 1.**
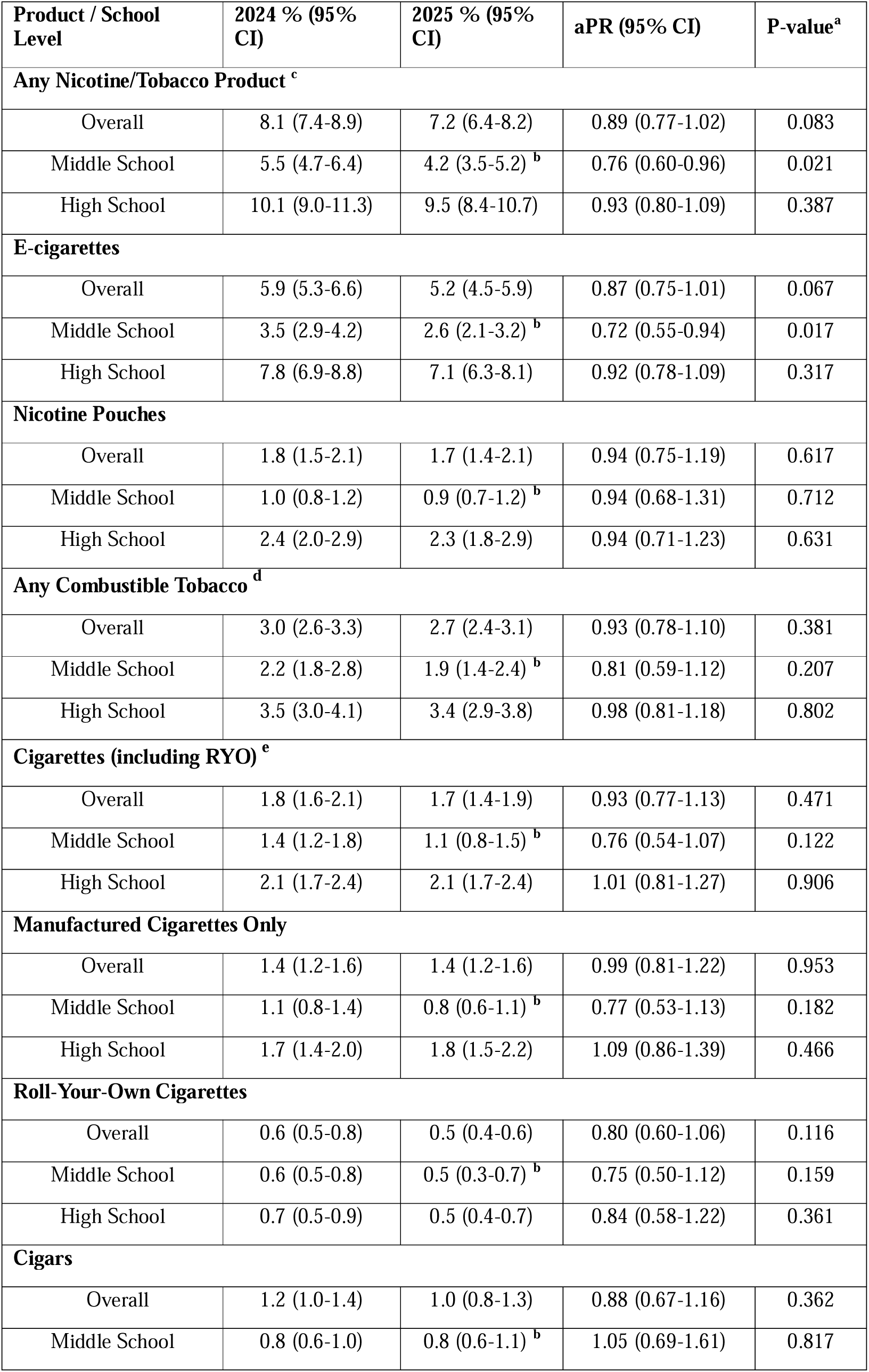
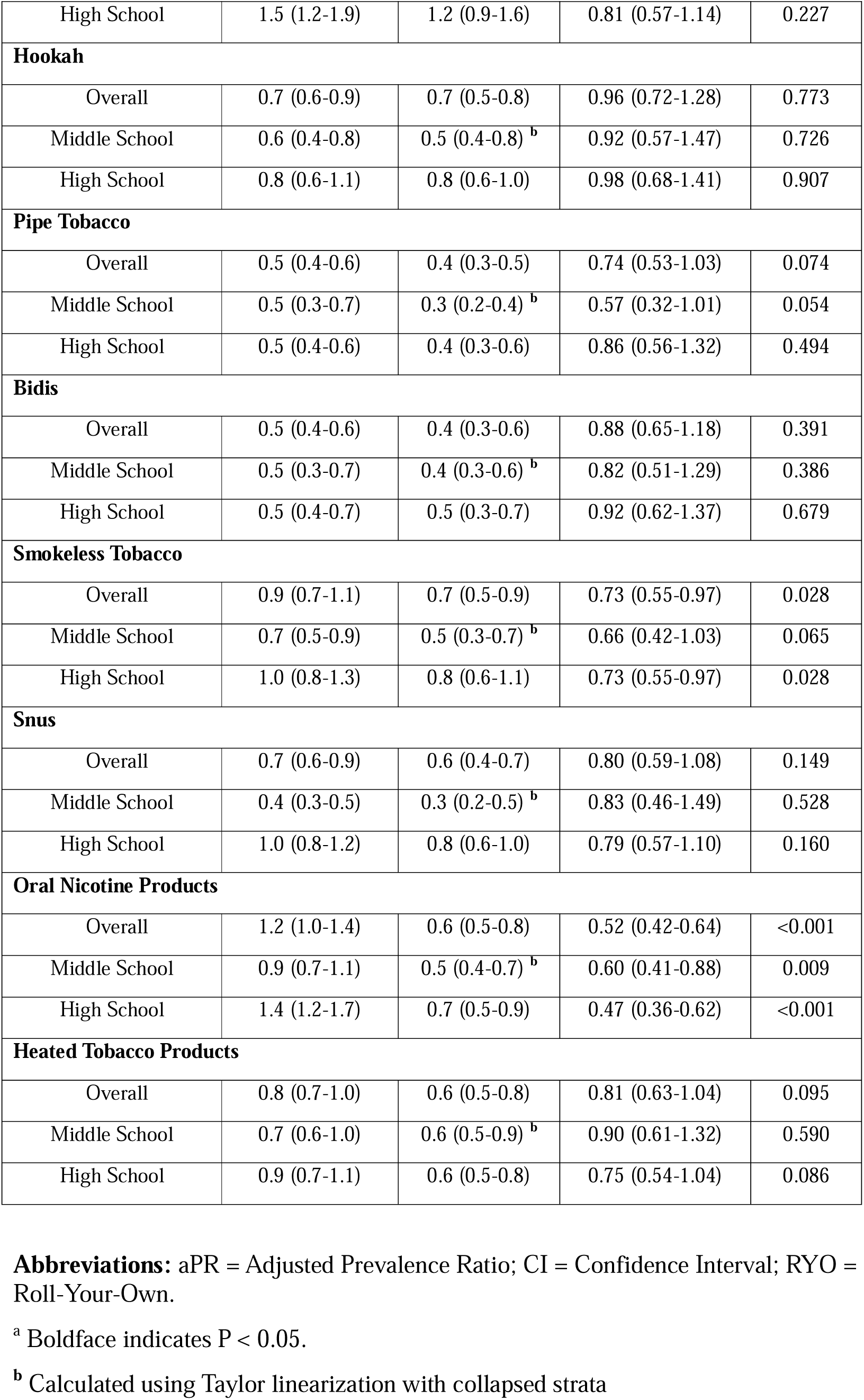
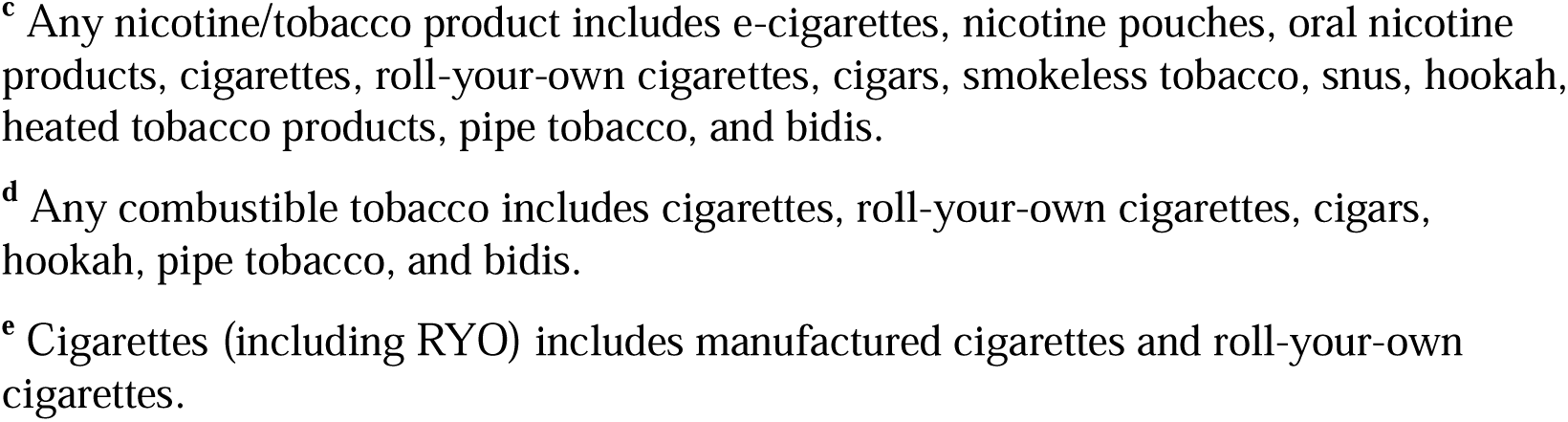
Estimated Prevalence of Past 30-Day Nicotine/Tobacco Product Use among U.S. Middle and High School Students, 2024–2025.

| Product / School Level | 2024 % (95% CI) | 2025 % (95% CI) | aPR (95% CI) | P-value<sup>a</sup> |
| --- | --- | --- | --- | --- |
| **Any Nicotine/Tobacco Product<sup>c</sup>** |  |  |  |  |
| Overall | 8.1 (7.4-8.9) | 7.2 (6.4-8.2) | 0.89 (0.77-1.02) | 0.083 |
| Middle School | 5.5 (4.7-6.4) | 4.2 (3.5-5.2)<sup>b</sup> | 0.76 (0.60-0.96) | 0.021 |
| High School | 10.1 (9.0-11.3) | 9.5 (8.4-10.7) | 0.93 (0.80-1.09) | 0.387 |
| **E-cigarettes** |  |  |  |  |
| Overall | 5.9 (5.3-6.6) | 5.2 (4.5-5.9) | 0.87 (0.75-1.01) | 0.067 |
| Middle School | 3.5 (2.9-4.2) | 2.6 (2.1-3.2)<sup>b</sup> | 0.72 (0.55-0.94) | 0.017 |
| High School | 7.8 (6.9-8.8) | 7.1 (6.3-8.1) | 0.92 (0.78-1.09) | 0.317 |
| **Nicotine Pouches** |  |  |  |  |
| Overall | 1.8 (1.5-2.1) | 1.7 (1.4-2.1) | 0.94 (0.75-1.19) | 0.617 |
| Middle School | 1.0 (0.8-1.2) | 0.9 (0.7-1.2)<sup>b</sup> | 0.94 (0.68-1.31) | 0.712 |
| High School | 2.4 (2.0-2.9) | 2.3 (1.8-2.9) | 0.94 (0.71-1.23) | 0.631 |
| **Any Combustible Tobacco<sup>d</sup>** |  |  |  |  |
| Overall | 3.0 (2.6-3.3) | 2.7 (2.4-3.1) | 0.93 (0.78-1.10) | 0.381 |
| Middle School | 2.2 (1.8-2.8) | 1.9 (1.4-2.4)<sup>b</sup> | 0.81 (0.59-1.12) | 0.207 |
| High School | 3.5 (3.0-4.1) | 3.4 (2.9-3.8) | 0.98 (0.81-1.18) | 0.802 |
| **Cigarettes (including RYO)<sup>e</sup>** |  |  |  |  |
| Overall | 1.8 (1.6-2.1) | 1.7 (1.4-1.9) | 0.93 (0.77-1.13) | 0.471 |
| Middle School | 1.4 (1.2-1.8) | 1.1 (0.8-1.5)<sup>b</sup> | 0.76 (0.54-1.07) | 0.122 |
| High School | 2.1 (1.7-2.4) | 2.1 (1.7-2.4) | 1.01 (0.81-1.27) | 0.906 |
| **Manufactured Cigarettes Only** |  |  |  |  |
| Overall | 1.4 (1.2-1.6) | 1.4 (1.2-1.6) | 0.99 (0.81-1.22) | 0.953 |
| Middle School | 1.1 (0.8-1.4) | 0.8 (0.6-1.1)<sup>b</sup> | 0.77 (0.53-1.13) | 0.182 |
| High School | 1.7 (1.4-2.0) | 1.8 (1.5-2.2) | 1.09 (0.86-1.39) | 0.466 |
| **Roll-Your-Own Cigarettes** |  |  |  |  |
| Overall | 0.6 (0.5-0.8) | 0.5 (0.4-0.6) | 0.80 (0.60-1.06) | 0.116 |
| Middle School | 0.6 (0.5-0.8) | 0.5 (0.3-0.7)<sup>b</sup> | 0.75 (0.50-1.12) | 0.159 |
| High School | 0.7 (0.5-0.9) | 0.5 (0.4-0.7) | 0.84 (0.58-1.22) | 0.361 |
| **Cigars** |  |  |  |  |
| Overall | 1.2 (1.0-1.4) | 1.0 (0.8-1.3) | 0.88 (0.67-1.16) | 0.362 |
| Middle School | 0.8 (0.6-1.0) | 0.8 (0.6-1.1)<sup>b</sup> | 1.05 (0.69-1.61) | 0.817 |
| High School | 1.5 (1.2-1.9) | 1.2 (0.9-1.6) | 0.81 (0.57-1.14) | 0.227 |
| **Hookah** |  |  |  |  |
| Overall | 0.7 (0.6-0.9) | 0.7 (0.5-0.8) | 0.96 (0.72-1.28) | 0.773 |
| Middle School | 0.6 (0.4-0.8) | 0.5 (0.4-0.8)<sup>b</sup> | 0.92 (0.57-1.47) | 0.726 |
| High School | 0.8 (0.6-1.1) | 0.8 (0.6-1.0) | 0.98 (0.68-1.41) | 0.907 |
| **Pipe Tobacco** |  |  |  |  |
| Overall | 0.5 (0.4-0.6) | 0.4 (0.3-0.5) | 0.74 (0.53-1.03) | 0.074 |
| Middle School | 0.5 (0.3-0.7) | 0.3 (0.2-0.4)<sup>b</sup> | 0.57 (0.32-1.01) | 0.054 |
| High School | 0.5 (0.4-0.6) | 0.4 (0.3-0.6) | 0.86 (0.56-1.32) | 0.494 |
| **Bidis** |  |  |  |  |
| Overall | 0.5 (0.4-0.6) | 0.4 (0.3-0.6) | 0.88 (0.65-1.18) | 0.391 |
| Middle School | 0.5 (0.3-0.7) | 0.4 (0.3-0.6)<sup>b</sup> | 0.82 (0.51-1.29) | 0.386 |
| High School | 0.5 (0.4-0.7) | 0.5 (0.3-0.7) | 0.92 (0.62-1.37) | 0.679 |
| **Smokeless Tobacco** |  |  |  |  |
| Overall | 0.9 (0.7-1.1) | 0.7 (0.5-0.9) | 0.73 (0.55-0.97) | 0.028 |
| Middle School | 0.7 (0.5-0.9) | 0.5 (0.3-0.7)<sup>b</sup> | 0.66 (0.42-1.03) | 0.065 |
| High School | 1.0 (0.8-1.3) | 0.8 (0.6-1.1) | 0.73 (0.55-0.97) | 0.028 |
| **Snus** |  |  |  |  |
| Overall | 0.7 (0.6-0.9) | 0.6 (0.4-0.7) | 0.80 (0.59-1.08) | 0.149 |
| Middle School | 0.4 (0.3-0.5) | 0.3 (0.2-0.5)<sup>b</sup> | 0.83 (0.46-1.49) | 0.528 |
| High School | 1.0 (0.8-1.2) | 0.8 (0.6-1.0) | 0.79 (0.57-1.10) | 0.160 |
| **Oral Nicotine Products** |  |  |  |  |
| Overall | 1.2 (1.0-1.4) | 0.6 (0.5-0.8) | 0.52 (0.42-0.64) | <0.001 |
| Middle School | 0.9 (0.7-1.1) | 0.5 (0.4-0.7)<sup>b</sup> | 0.60 (0.41-0.88) | 0.009 |
| High School | 1.4 (1.2-1.7) | 0.7 (0.5-0.9) | 0.47 (0.36-0.62) | <0.001 |
| **Heated Tobacco Products** |  |  |  |  |
| Overall | 0.8 (0.7-1.0) | 0.6 (0.5-0.8) | 0.81 (0.63-1.04) | 0.095 |
| Middle School | 0.7 (0.6-1.0) | 0.6 (0.5-0.9)<sup>b</sup> | 0.90 (0.61-1.32) | 0.590 |
| High School | 0.9 (0.7-1.1) | 0.6 (0.5-0.8) | 0.75 (0.54-1.04) | 0.086 |

**Abbreviations:** aPR = Adjusted Prevalence Ratio; CI = Confidence Interval; RYO = Roll-Your-Own.
<sup>a</sup> Boldface indicates $P < 0.05$ .
<sup>b</sup> Calculated using Taylor linearization with collapsed strata
<sup>c</sup> Any nicotine/tobacco product includes e-cigarettes, nicotine pouches, oral nicotine products, cigarettes, roll-your-own cigarettes, cigars, smokeless tobacco, snus, hookah, heated tobacco products, pipe tobacco, and bidis.
<sup>d</sup> Any combustible tobacco includes cigarettes, roll-your-own cigarettes, cigars, hookah, pipe tobacco, and bidis.
<sup>e</sup> Cigarettes (including RYO) includes manufactured cigarettes and roll-your-own cigarettes.

By school level, 4.2% of middle school students and 9.5% of high school students reported past-30-day use of any product in 2025. Adjusted prevalence was lower in 2025 than 2024 among middle school students (aPR 0.76; 95% CI 0.60–0.96), while the change among high school students was smaller and the confidence interval included the null (aPR 0.93; 95% CI 0.80–1.09).

### Types of Products Used

E-cigarettes remained the most commonly used nicotine/tobacco product, while nicotine pouches and combustible tobacco were less prevalent (**Table 1**).

Overall, past 30-day e-cigarette use was 5.2% in 2025 (∼1.43 million students), down from 5.9% in 2024 (∼1.62 million). In 2025, prevalence was 2.6% among middle school students and 7.1% among high school students. Adjusted prevalence was lower in 2025 among middle school students (aPR 0.72; 95% CI 0.55–0.94), while among high school students the change was modest and compatible with no difference (aPR 0.92; 95% CI 0.78–1.09).

Past 30-day nicotine pouch use was 1.7% in 2025 (∼450,000 students), similar to 1.8% in 2024 (∼470,000). In 2025, prevalence was 0.9% among middle school students and 2.3% among high school students, with prevalence similar across years overall and by school level. Prevalence of nicotine pouch use remained higher among boys (2.4%) than girls (1.0%; **Supplementary Table 2**).

Among other non-combustible products, smokeless tobacco use declined modestly overall from 0.9% in 2024 to 0.7% in 2025 (aPR 0.73; 95% CI, 0.55-0.97), whereas oral nicotine product use declined more markedly from 1.2% to 0.6% (aPR 0.52; 95% CI, 0.42-0.64). Oral nicotine product use was lower in 2025 in both middle school students (0.9% to 0.5%; aPR 0.60; 95% CI 0.41–0.88) and high school students (1.4% to 0.7%; aPR 0.47; 95% CI 0.36–0.62).

Past 30-day use of any combustible tobacco was 2.7% in 2025, compared with 3.0% in 2024. In 2025, prevalence was 1.9% among middle school students and 3.4% among high school students. Cigarettes, including roll-your-own cigarettes, were the most commonly reported combustible product, with prevalence of 1.7% overall; 1.1% among middle school students and 2.1% among high school students. Among other combustible products, cigars (1.0%), hookah (0.7%), pipe tobacco (0.4%), and bidis (0.4%) were each used by no more than 1% of students in 2025, with prevalence similar to 2024. Heated tobacco products (0.6%) and snus (0.6%) also remained uncommon in 2025, with prevalence comparable across years. Complete prevalence estimates for all individual products are presented in **Table 1**, with sex-specific estimates in **Supplementary Table 2.**

### E-Cigarette Use Characteristics

In 2025, 40.7% of past 30-day e-cigarette users reported frequent use (≥20 of the past 30 days) and 27.0% reported daily use, which were comparable to 2024 (**Table 2**).

**Table 2.**
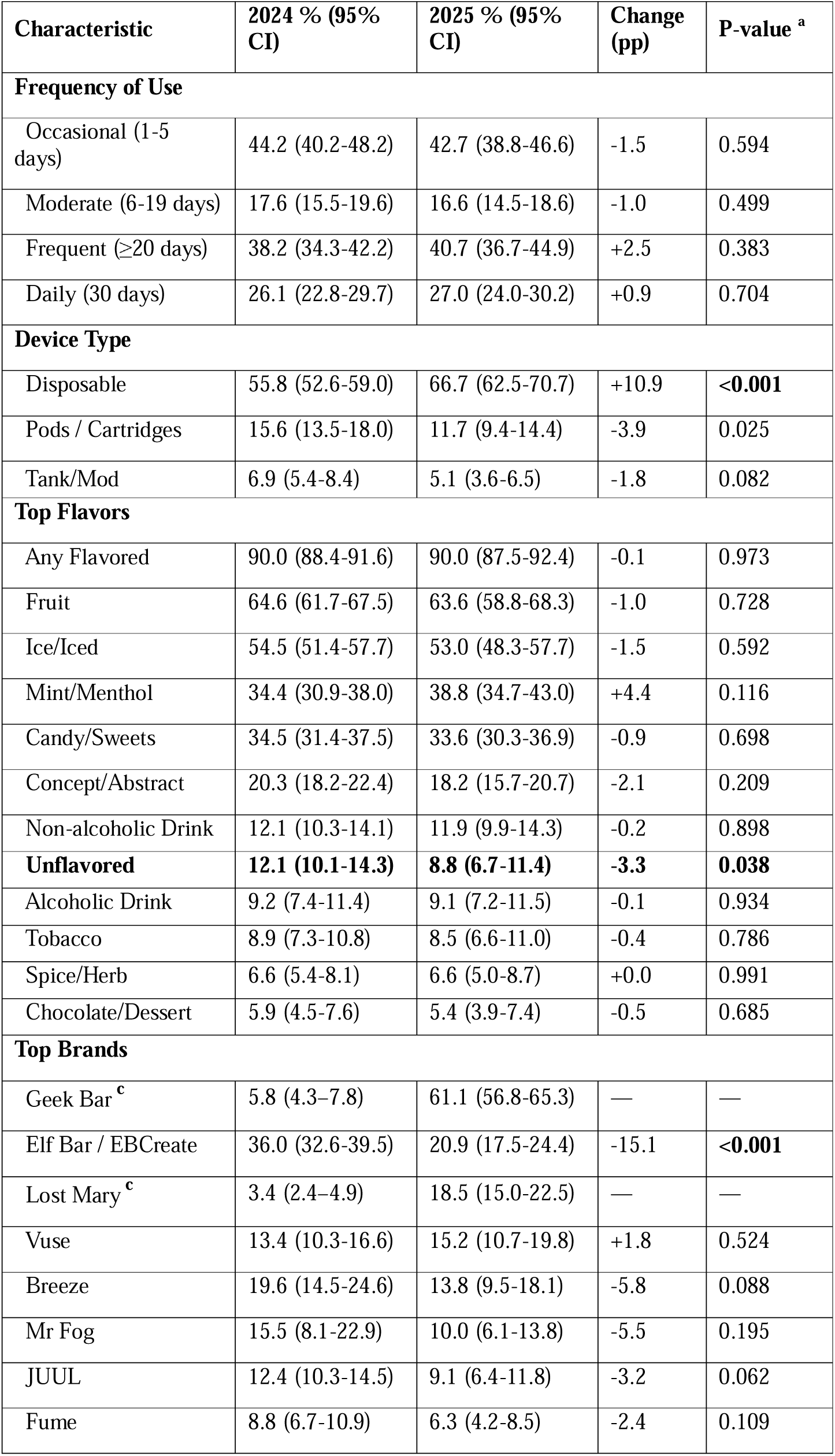
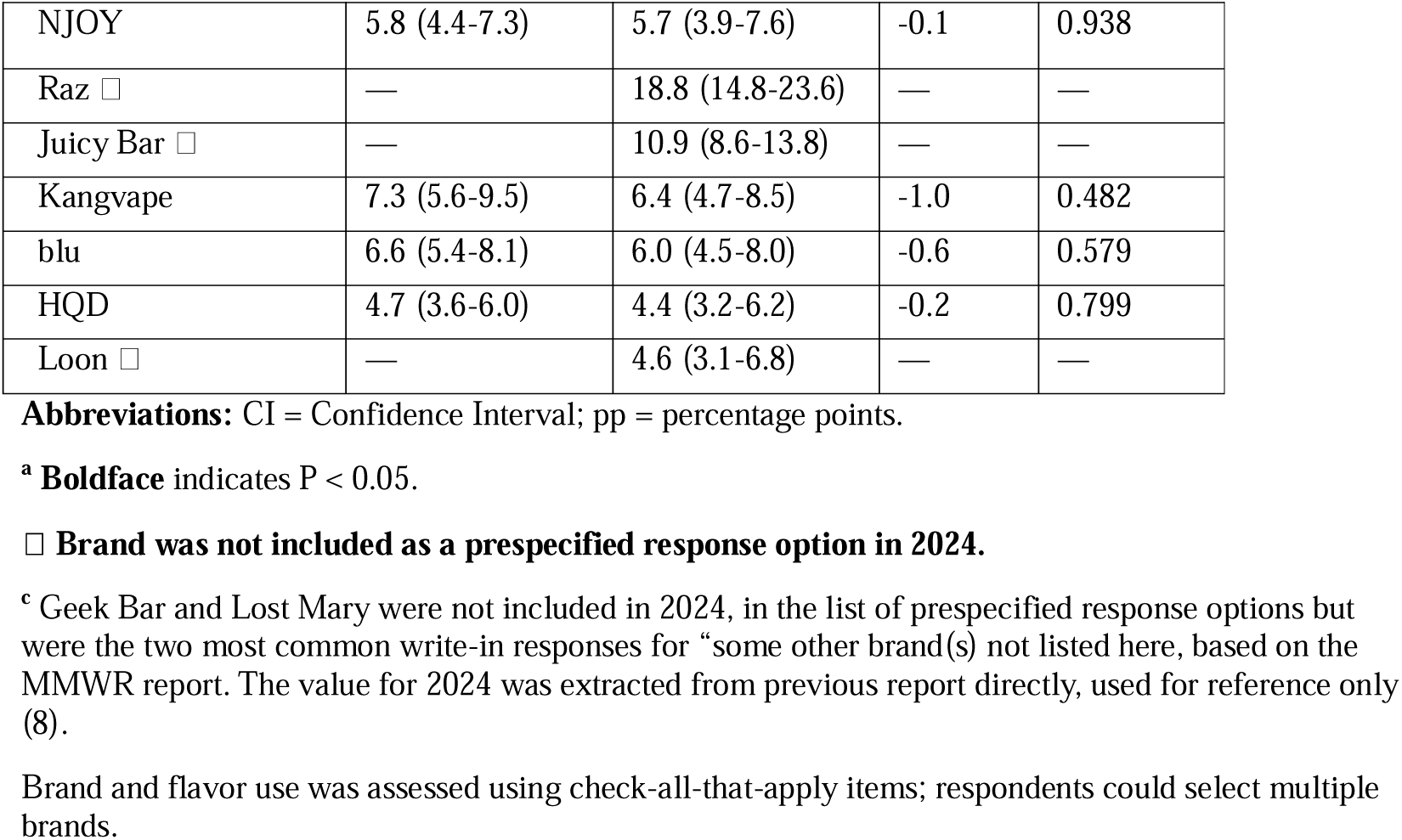
Characteristics and Preferences among Past 30-Day E-Cigarette Users, 2024–2025.

Device preferences shifted notably between survey years. Use of disposable e-cigarettes increased from 55.8% in 2024 to 66.7% in 2025 (P<0.001), while use of pod/cartridge-based devices declined from 15.6% to 11.7%) (P=0.022).

Brand preferences also shifted. In 2025, Geek Bar was the most commonly reported brand (61.1%), although it was not included as a prespecified response option in 2024. Elf Bar/EBCreate declined from 36.0% in 2024 to 20.9% in 2025 (P<0.001). Other popular brands in 2025 included Lost Mary (18.5%), Vuse (15.2%), Breeze (13.8%), and JUUL (9.1%).

Flavored e-cigarette use remained high, with 90.0% of users reporting any flavored product in 2025, unchanged from 2024. In contrast, unflavored e-cigarette use declined from 12.1% in 2024 to 8.8% in 2025 (P=0.038). Fruit remained the most commonly used flavor (63.6%), followed by ice/iced (53.0%), mint/menthol (38.8%), and candy/sweets (33.6%) in 2025. Concept or abstract flavors were less frequently used (18.2%). Tobacco-flavored e-cigarette use was reported by 8.5% of users, comparable to 2024.

### Nicotine Pouch Use Characteristics

In 2025, 26.0% of past 30-day nicotine pouch users reported frequent use (≥20 of the past 30 days) and 17.3% reported daily use, both lower than in 2024 (29.3% and 22.3%, respectively), although the confidence intervals for the changes were wide and included no difference **(Table 3**).

**Table 3.** Characteristics and Preferences among Past 30-Day Nicotine Pouch Users, 2024–2025.

| Characteristic | 2024 % (95% CI) | 2025 % (95% CI) | Change (pp) | P-value <sup>a</sup> |
| --- | --- | --- | --- | --- |
| **Frequency of Use** |  |  |  |  |
| Occasional (1-5 days) | 53.9 (48.0-59.8) | 53.0 (44.9-61.1) | -0.9 | 0.858 |
| Moderate (6-19 days) | 16.8 (12.7-21.0) | 21.0 (14.5-27.5) | 4.1 | 0.292 |
| Frequent (≥20 days) | 29.3 (24.6-34.4) | 26.0 (22.0-30.5) | -3.2 | 0.325 |
| Daily (30 days) | 22.3 (18.3-27.0) | 17.3 (14.0-21.2) | -5 | 0.081 |
| **Top Flavors** |  |  |  |  |
| Any Flavored | 83.6 (79.6-87.6) | 88.0 (84.2-91.8) | +4.5 | 0.114 |
| Mint/Menthol | 64.7 (58.7-70.7) | 70.3 (64.9-75.7) | 5.6 | 0.172 |
| Fruit | 23.5 (18.4-28.6) | 25.8 (19.6-31.9) | 2.3 | 0.574 |
| Ice/Iced | 23.2 (19.5-27.0) | 17.9 (13.3-22.6) | -5.3 | 0.084 |
| Concept/Abstract | 11.5 (8.1-15.0) | 11.2 (7.7-14.7) | -0.3 | 0.896 |
| Candy/Sweets | 10.4 (7.7-13.0) | 8.6 (5.1-12.1) | -1.8 | 0.438 |
| Spice/Herb | 11.1 (8.1-15.0) | 10.3 (7.2-14.6) | -0.7 | 0.770 |
| Tobacco | 8.7 (6.5-11.7) | 4.7 (3.1-7.0) | -4.1 | 0.012 |
| Chocolate/Dessert | 8.4 (6.1-11.5) | 5.7 (3.1-10.0) | -2.8 | 0.199 |
| Non-alcoholic Drink | 7.7 (5.4-10.8) | 9.7 (6.1-15.3) | +2.0 | 0.443 |
| Alcoholic Drink | 6.8 (4.6-10.0) | 5.3 (3.4-8.1) | -1.6 | 0.374 |
| Unflavored | 14.3 (10.7-18.9) | 6.4 (3.9-10.2) | -8.0 | 0.002 |
| **Top Brands** |  |  |  |  |
| ZYN | 68.3 (62.3-74.2) | 69.4 (62.5-76.3) | 1.1 | 0.808 |
| on! | 14.1 (10.6-17.5) | 14.4 (10.7-18.1) | 0.3 | 0.894 |
| VELO | 10.6 (7.8-13.3) | 12.4 (3.9-20.9) | 1.9 | 0.684 |
| Rogue | 13.5 (10.0-16.9) | 12.3 (9.1-15.5) | -1.2 | 0.625 |
| Juice Head | 9.6 (7.1-12.0) | 7.3 (3.8-10.8) | -2.3 | 0.295 |
| FRE | 9.3 (6.4-12.3) | 7.1 (3.9-10.2) | -2.3 | 0.299 |
| 2ONE/ZONE ☐ | 7.0 (5.0-9.8) | 10.5 (6.0-17.8) | +3.5 | 0.266 |
| Grizzly ☐ | — | 9.2 (6.5-12.8) | — | — |
| Black Buffalo ☐ | — | 4.0 (2.5-6.4) | — | — |
| ZEO ☐ | — | 3.9 (2.0-7.6) | — | — |

**Abbreviations:** CI = Confidence Interval; pp = percentage points.
☐ Boldface indicates P < 0.05.
☐ Brand was not included as a prespecified response option in 2024.
☐ Listed as "2ONE" in 2024 and "ZONE" in 2025; treated as the same brand.
Brand and flavor use was assessed using check-all-that-apply items; respondents could select multiple brands.

Flavored nicotine pouch use remained high, reported by 88.0% of users in 2025, compared with 83.6% in 2024. In contrast, unflavored pouch use declined from 14.3% in 2024 to 6.4% in 2025 (P=0.002). Mint/menthol was the most commonly used flavor (70.3%), followed by fruit (25.8%). Other flavors, including ice/iced, candy/sweets, and concept/abstract, were less frequently used. Use of tobacco-flavored pouches also declined, from 8.7% in 2024 to 4.7% in 2025 (P=0.012).

Brand preferences were relatively stable. ZYN remained the dominant brand at 69.4% in 2025, similar to 68.3% in 2024. Other brands were reported by smaller proportions of users, with no notable changes between survey years.

## Discussion

In this repeated cross-sectional study of nationally representative U.S. middle and high school students, overall past 30-day nicotine and tobacco product use was lower in 2025 than 2024, continuing the downward trajectory observed since 2019 (5, 11). Cigarette smoking prevalence remained at 1.7%, and overall combustible tobacco use remained low at 2.7%. E-cigarettes remained the most commonly used product, with a notable shift in brand dominance to Geek Bar and an increase in disposable device use. Nicotine pouch use was stable at approximately 1.7%, with ZYN reported by nearly 70% of pouch users.

The dominance of Geek Bar among e-cigarette users in 2025, alongside the decline of Elf Bar/EBCreate, illustrates the dynamic reorganization of the youth e-cigarette market. Shifts in brand popularity did not reduce overall e-cigarette use, highlighting how youth may readily switch between products rather than stop using nicotine. The continued growth of disposable e-cigarettes is consistent with international trends. In England, the rapid rise of disposable devices was associated with increases in vaping among young adults, including those who had never regularly smoked (12, 13). These concerns, along with environmental ones, prompted the United Kingdom to legislate a ban on disposable vapes (14). When the United Kingdom legislated a ban on disposable vapes, the proportion of vapers using disposables declined, but young adult vapers (aged 16-24) shifted to refillable and rechargeable devices rather than stopping (15). This pattern of product substitution, in which users switch between formats rather than cease use, is consistent with the brand-switching observed in our US data, and suggests that the impact of product-specific regulations on overall nicotine use among youth requires careful monitoring.

The ongoing decline in combustible tobacco use among youth represents a sustained public health achievement. Cigarette prevalence remained at 1.7%, the lowest level ever recorded by the NYTS, and other combustible products, including cigars, hookah, and pipe tobacco, were each used by no more than 1% of students, with prevalence comparable across years (5). In relation to newer products, prevalence of nicotine pouch use remained stable across 2024 and 2025, following a period of rapid growth in earlier NYTS waves, during which prevalence increased rapidly while the number of users who had never used other tobacco or nicotine products increased substantially (16). The composition of pouch use has shifted: unflavored or tobacco-flavored pouch use declined while flavored use, predominantly mint or menthol, remained high. Evidence from e-cigarettes suggests that unflavored and tobacco-flavored products are more commonly used by existing tobacco users (17). If this pattern extends to nicotine pouches, the shift toward flavored products may signal a user base that is increasingly composed of individuals without prior tobacco experience. Although cross-sectional data cannot distinguish whether stable prevalence reflects continued use by the same individuals or turnover in the user population, a key question is whether pouches are substituting for more harmful tobacco products or instead attracting young people who would otherwise not use nicotine (18). This distinction has important implications for the regulation of oral nicotine pouches. Oral nicotine product use (including lozenges, gums, and dissolvable products) also declined notably. This pattern is consistent with broader market trends, showing that the rapid growth of nicotine pouches has coincided with considerable declines in sales of other oral tobacco and nicotine products (19).

Declines in nicotine and tobacco use were concentrated among middle school students for both any tobacco product use and e-cigarette use, while high school prevalence showed less evidence of change. This age-differential pattern is consistent with the possibility that existing tobacco control measures, such as minimum legal sales age requirements and school-based prevention campaigns, may be more effective at deterring initiation among younger adolescents (5, 20, 21). It may also reflect that high school prevalence has already fallen substantially from earlier peaks (5) or that the available data were insufficient to detect a change within broader influences on youth behavior, as the confidence interval included the possibility of notable year-on-year variation.

This study has several limitations. First, the 2025 NYTS response rate was 29.7%, lower than the 33.4% in 2024, which may affect the representativeness of estimates despite the application of survey weights (5, 6). Second, cross-year comparisons of brand prevalence should be interpreted cautiously, as several brands, like Geek Bar and Lost Mary, were not included as prespecified response options in 2024 (8). Third, all measures were self-reported and may be subject to recall or social desirability bias. Fourth, complete-case analysis was used, and missingness on some variables may have introduced bias if data were not missing completely at random.

These findings indicate that overall youth nicotine and tobacco use continues to decline, and the product landscape is evolving rapidly. The speed of brand turnover in the e-cigarette market highlights the challenges of regulating a rapidly evolving product marketplace. The consolidation of flavored nicotine pouch use, alongside declines in use of unflavored and tobacco-flavored products, requires further investigation into whether the composition of the youth user base is changing over time. Continued product-specific surveillance will be essential to inform timely regulatory and public health responses to these rapidly changing markets.

## Data Availability

All data produced are available online at https://www.fda.gov/tobacco-products/youth-and-tobacco/national-youth-tobacco-survey-nyts

## Reference

1. Warner KE. Kids No Longer Smoke Cigarettes. Why Aren’t We Celebrating? Am J Public Health. 2024;114(11):1191–4.

2. Birdsey J. Tobacco product use among US middle and high school students—National Youth Tobacco Survey, 2023. MMWR Morbidity and mortality weekly report. 2023;72.

3. Gentzke AS. Tobacco product use and associated factors among middle and high school students—National Youth Tobacco Survey, United States, 2021. MMWR Surveillance Summaries. 2022;71.

4. US Food and Drug Administration. National Youth Tobacco Survey (NYTS), Silver Spring, MD: FDA Tobacco Products; 2026 [cited 2026 March 12]. Available from: https://www.fda.gov/tobacco-products/youth-and-tobacco/national-youth-tobacco-survey-nyts.

5. Jamal A, Park-Lee E, Birdsey J, West A, Cornelius M, Cooper MR, et al. Tobacco Product Use Among Middle and High School Students - National Youth Tobacco Survey, United States, 2024. MMWR Morb Mortal Wkly Rep. 2024;73(41):917–24.

6. Products CfT. 2025 National Youth Tobacco Survey: Methodology Report.; 2025.

7. Health OoSa. 2024 NATIONAL YOUTH TOBACCO SURVEY: METHODOLOGY REPORT. 2024.

8. Park-Lee E, Jamal A, Cowan H, Sawdey MD, Cooper MR, Birdsey J, et al. Notes from the Field: E-Cigarette and Nicotine Pouch Use Among Middle and High School Students - United States, 2024. MMWR Morb Mortal Wkly Rep. 2024;73(35):774–8.

9. West BT, Berglund P, Heeringa SG. A Closer Examination of Subpopulation Analysis of Complex-Sample Survey Data. The Stata Journal. 2008;8(4):520–31.

10. Rust KF, Rao JN. Variance estimation for complex surveys using replication techniques. Stat Methods Med Res. 1996;5(3):283–310.

11. Jackson SE, Brown J, Tattan-Birch H, Jarvis MJ. Changing patterns of nicotine product use and nicotine dependence among United States high-school students: The National Youth Tobacco Survey, 2014–2023. Addiction. 2025;120(11):2215–22.

12. Tattan-Birch H, Brown J, Shahab L, Beard E, Jackson SE. Trends in vaping and smoking following the rise of disposable e-cigarettes: a repeat cross-sectional study in England between 2016 and 2023. Lancet Reg Health Eur. 2024;42:100924.

13. Jackson SE, Shahab L, Buss V, Tattan-Birch H, Cox S, Taylor E, Brown J. The changing face of nicotine use in England: Age-specific annual trends, 2014 to 2024. Addiction. 2026;121(3):549–63.

14. Department for Environment FRA, Department of Health and Social Care,. Single-use vapes banned from 1 June 2025: GOV.UK; 2025 [Available from: https://www.gov.uk/government/news/single-use-vapes-banned-from-1-june-2025.

15. Jackson SE, Shahab L, Tattan-Birch H, Buss V, Brown J. Changes in vaping trends since the announcement of an impending ban on disposable vapes: A population study in Great Britain. Addiction. 2025;120(9):1876–83.

16. Sun H, Tattan-Birch H, Oldham M, Cox S, Jackson SE. Oral Nicotine Pouch Use Among U.S. Middle and High School Students, 2021-2023. medRxiv. 2026:2026.01.28.26345040.

17. Gades MS, Alcheva A, Riegelman AL, Hatsukami DK. The Role of Nicotine and Flavor in the Abuse Potential and Appeal of Electronic Cigarettes for Adult Current and Former Cigarette and Electronic Cigarette Users: A Systematic Review. Nicotine Tob Res. 2022;24(9):1332–43.

18. Travis N, Warner KE, Goniewicz ML, Oh H, Ranganathan R, Meza R, et al. The Potential Impact of Oral Nicotine Pouches on Public Health: A Scoping Review. Nicotine Tob Res. 2025;27(4):598–610.

19. Hrywna M, Wackowski OA, Robichaud MO, Talbot EM, Barnwell PV, Delnevo CD. Oral Tobacco and Nicotine Marketplace Trends Since the Tobacco Control Act. JAMA Network Open. 2025;8(10):e2540747-e.

20. MacMonegle A, Nguyen Zarndt A, Wang Y, Bennett M, Malo V, Pitzer L, et al. The Impact of “The Real Cost” on E-cigarette Initiation Among U.S. Youth. American Journal of Preventive Medicine. 2025:107614.

21. Centers for Disease Control and Prevention OoSaH. Summary of Scientific Evidence: Raising the Minimum Legal Sales Age for Tobacco Products. 2021.

