## Supplementary table 1 and 2 for "Nicotine and tobacco product use among US middle and high school students, 2024-25"

**Supplementary Table 1.** Sensitivity analyses comparing variance estimation methods for 2025 middle school subgroup prevalence estimates of past 30-day nicotine/tobacco product use.

| Outcome | Taylor (original) | Taylor (collapsed) | JKn (original) | JKn (collapsed) | Bootstrap (original) | Bootstrap (collapsed) | Fay's BRR | Weights only |
| --- | --- | --- | --- | --- | --- | --- | --- | --- |
| Any  tobacco | NA | 3.93 (3.21-4.80) | 3.93 (3.21-4.81) | 3.93 (3.20-4.82) | 3.93 (3.21-4.81) | 3.93 (3.21-4.80) | 3.92 (3.28-4.69) | 3.93 (3.55-4.34) |
| E-cigarette | NA | 2.57 (2.08-3.17) | 2.57 (2.08-3.17) | 2.57 (2.07-3.18) | 2.57 (2.06-3.19) | 2.57 (2.08-3.17) | 2.56 (2.07-3.16) | 2.57 (2.27-2.90) |
| Nicotine Pouches | NA | 0.93 (0.73-1.18) | 0.93 (0.72-1.19) | 0.93 (0.72-1.19) | 0.93 (0.73-1.18) | 0.93 (0.72-1.19) | 0.92 (0.71-1.19) | 0.93 (0.75-1.15) |
| Combustible  Tobacco | NA | 1.86 (1.41-2.44) | 1.86 (1.41-2.45) | 1.86 (1.41-2.46) | 1.86 (1.41-2.44) | 1.86 (1.40-2.47) | 1.85 (1.48-2.31) | 1.86 (1.61-2.15) |
| Cigarettes | NA | 0.82 (0.61-1.10) | 0.82 (0.61-1.10) | 0.82 (0.60-1.11) | 0.82 (0.61-1.10) | 0.82 (0.60-1.11) | 0.81 (0.65-1.02) | 0.82 (0.65-1.03) |
| Roll-your-own Cigarettes | NA | 0.47 (0.34-0.66) | 0.47 (0.34-0.66) | 0.47 (0.34-0.66) | 0.47 (0.34-0.66) | 0.47 (0.33-0.68) | 0.47 (0.34-0.65) | 0.47 (0.36-0.63) |
| Any Cigarettes | NA | 1.12 (0.84-1.48) | 1.12 (0.84-1.48) | 1.12 (0.84-1.49) | 1.12 (0.85-1.48) | 1.12 (0.84-1.50) | 1.11 (0.91-1.36) | 1.12 (0.92-1.35) |
| cigar | NA | 0.79 (0.56-1.12) | 0.79 (0.56-1.12) | 0.79 (0.56-1.13) | 0.79 (0.56-1.13) | 0.79 (0.57-1.11) | 0.78 (0.57-1.08) | 0.79 (0.63-1.00) |
| Heated tobacco product | NA | 0.64 (0.48-0.86) | 0.64 (0.48-0.87) | 0.64 (0.48-0.87) | 0.64 (0.48-0.87) | 0.64 (0.48-0.86) | 0.64 (0.50-0.81) | 0.64 (0.50-0.83) |
| Hookah | NA | 0.54 (0.36-0.80) | 0.54 (0.35-0.81) | 0.54 (0.35-0.81) | 0.54 (0.35-0.81) | 0.54 (0.36-0.79) | 0.53 (0.35-0.80) | 0.54 (0.41-0.70) |
| Smokeless Tobacco | NA | 0.47 (0.33-0.65) | 0.47 (0.33-0.65) | 0.47 (0.33-0.65) | 0.47 (0.33-0.65) | 0.47 (0.33-0.66) | 0.46 (0.34-0.62) | 0.47 (0.35-0.62) |
| Snus | NA | 0.31 (0.19-0.51) | 0.31 (0.19-0.51) | 0.31 (0.19-0.52) | 0.31 (0.18-0.52) | 0.31 (0.19-0.52) | 0.30 (0.21-0.44) | 0.31 (0.22-0.44) |
| Oral nicotine products | NA | 0.55 (0.41-0.73) | 0.55 (0.40-0.73) | 0.55 (0.40-0.74) | 0.55 (0.40-0.73) | 0.55 (0.41-0.73) | 0.54 (0.42-0.69) | 0.55 (0.41-0.73) |
| Pipe | NA | 0.28 (0.18-0.44) | 0.28 (0.18-0.44) | 0.28 (0.17-0.45) | 0.28 (0.17-0.46) | 0.28 (0.17-0.45) | 0.27 (0.18-0.40) | 0.28 (0.19-0.41) |
| Bidis | NA | 0.40 (0.28-0.58) | 0.40 (0.28-0.59) | 0.40 (0.28-0.59) | 0.40 (0.28-0.58) | 0.40 (0.28-0.59) | 0.40 (0.27-0.59) | 0.40 (0.30-0.55) |

Prevalence estimates (%) and 95% confidence intervals are shown for each outcome across seven variance estimation approaches: Taylor linearization (original and collapsed strata), Jackknife (original and collapsed), subbootstrap (original and collapsed), Fay's balanced repeated replication (BRR), and weights-only estimation. "NA" indicates non-estimable confidence intervals due to singleton primary sampling unit strata. Adjacent strata were collapsed prior to variance estimation to address singleton PSUs in the 2025 middle school subgroup.

**Supplementary Table 2.** Estimated Prevalence of Past 30-Day Nicotine/Tobacco Product Use stratified by sex among U.S. Middle and High School Students, 2024–2025

| **Product / School Level** | **2024 % (95% CI)** | **2025 % (95% CI)** | **aPR (95% CI)** | **P-value^a^** |
| --- | --- | --- | --- | --- |
| **Any Nicotine/Tobacco Product ^c^** | | | | |
| Male | 8.5 (7.6-9.5) | 7.5 (6.4-8.5) | 0.87 (0.74-1.02) | 0.327 |
| Female | 7.7 (6.8-8.5) | 7.1 (6.1-8.0) | 0.91 (0.77-1.06) | 0.147 |
| **E-cigarettes** | | | | |
| Male | 5.8 (5.0-6.5) | 4.9 (4.1-5.7) | 0.85 (0.71-1.01) | 0.062 |
| Female | 6.1 (5.3-6.9) | 5.5 (4.7-6.3) | 0.90 (0.75-1.07) | 0.222 |
| **Nicotine Pouches** | | | | |
| Male | 2.7 (2.1-3.2) | 2.4 (1.9-3.0) | 0.89 (0.68-1.17) | 0.410 |
| Female | 0.8 (0.7-1.0) | 1.0 (0.7-1.2) | 1.10 (0.82-1.49) | 0.521 |
| **Any Combustible Tobacco ^d^** | | | | |
| Male | 3.3 (2.9-3.8) | 2.9 (2.4-3.4) | 0.88 (0.72-1.08) | 0.222 |
| Female | 2.5 (2.1-3.0) | 2.5 (2.1-2.9) | 0.99 (0.79-1.24) | 0.922 |
| **Cigarettes (including RYO) ^e^** | | | | |
| Male | 2.0 (1.7-2.4) | 1.7 (1.4-2.1) | 0.87 (0.68-1.10) | 0.237 |
| Female | 1.5 (1.3-1.8) | 1.6 (1.3-1.9) | 1.02 (0.80-1.30) | 0.872 |
| **Manufactured Cigarettes Only** | | | | |
| Male | 1.6 (1.3-1.9) | 1.5 (1.2-1.8) | 0.94 (0.73-1.22) | 0.648 |
| Female | 1.2 (0.9-1.4) | 1.3 (1.0-1.5) | 1.07 (0.81-1.41) | 0.633 |
| **Roll-Your-Own Cigarettes** | | | | |
| Male | 0.7 (0.5-0.9) | 0.5 (0.4-0.6) | 0.70 (0.48-1.01) | 0.059 |
| Female | 0.6 (0.4-0.8) | 0.5 (0.4-0.7) | 0.92 (0.62-1.38) | 0.700 |
| **Cigars** | | | | |
| Male | 1.5 (1.2-1.8) | 1.3 (1.0-1.7) | 0.89 (0.64-1.24) | 0.504 |
| Female | 0.9 (0.6-1.1) | 0.7 (0.5-1.0) | 0.85 (0.58-1.25) | 0.405 |
| **Hookah** | | | | |
| Male | 0.7 (0.5-0.9) | 0.6 (0.5-0.8) | 0.93 (0.66-1.32) | 0.694 |
| Female | 0.7 (0.5-0.9) | 0.7 (0.5-0.9) | 0.98 (0.67-1.44) | 0.933 |
| **Pipe Tobacco** | | | | |
| Male | 0.6 (0.4-0.7) | 0.4 (0.3-0.5) | 0.74 (0.49-1.10) | 0.138 |
| Female | 0.4 (0.3-0.5) | 0.3 (0.2-0.4) | 0.74 (0.46-1.21) | 0.230 |
| **Bidis** | | | | |
| Male | 0.6 (0.5-0.8) | 0.5 (0.4-0.7) | 0.86 (0.60-1.23) | 0.417 |
| Female | 0.4 (0.3-0.5) | 0.3 (0.2-0.5) | 0.91 (0.55-1.48) | 0.692 |
| **Smokeless Tobacco** | | | | |
| Male | 1.3 (1.0-1.6) | 1.0 (0.7-1.3) | 0.79 (0.58-1.08) | 0.145 |
| Female | 0.5 (0.4-0.6) | 0.3 (0.2-0.4) | 0.56 (0.37-0.86) | **0.007** |
| **Snus** | | | | |
| Male | 1.0 (0.8-1.2) | 0.9 (0.7-1.1) | 0.92 (0.67-1.28) | 0.629 |
| Female | 0.4 (0.3-0.5) | 0.2 (0.1-0.3) | 0.51 (0.32-0.82) | **0.005** |
| **Oral Nicotine Products** | | | | |
| Male | 1.5 (1.2-1.7) | 0.8 (0.6-0.9) | 0.51 (0.39-0.67) | <0.001 |
| Female | 0.9 (0.7-1.1) | 0.5 (0.3-0.6) | 0.52 (0.37-0.74) | <0.001 |
| **Heated Tobacco Products** | | | | |
| Male | 0.9 (0.7-1.1) | 0.8 (0.6-0.9) | 0.87 (0.64-1.18) | 0.367 |
| Female | 0.7 (0.5-0.9) | 0.5 (0.4-0.7) | 0.73 (0.49-1.09) | 0.127 |

**Abbreviations:** aPR = Adjusted Prevalence Ratio; CI = Confidence Interval; RYO = Roll-Your-Own.

^a^ Boldface indicates P < 0.05.

**^b^** calculated using Taylor linearization with collapsed strata

**^c^** Any nicotine/tobacco product includes e-cigarettes, nicotine pouches, oral nicotine products, cigarettes, roll-your-own cigarettes, cigars, smokeless tobacco, snus, hookah, heated tobacco products, pipe tobacco, and bidis.

**^d^** Any combustible tobacco includes cigarettes, roll-your-own cigarettes, cigars, hookah, pipe tobacco, and bidis.

**^e^** Cigarettes (including RYO) includes manufactured cigarettes and roll-your-own cigarettes.
